# Evaluating cranial electrotherapy stimulation for anxiety associated with breathlessness in palliative care: a mixed-methods feasibility study

**DOI:** 10.64898/2026.08.22.26361094

**Authors:** Lucy Bleazard, Sarah R Copping, Sarah Booth, Laura J Gray, Christina Faull, Kate Walker, Chris Griffiths, David Wenzel

## Abstract

**Objectives:** To explore the acceptability and tolerability of cranial electrotherapy stimulation (CES) using Alpha-Stim AID as a potential intervention for anxiety associated with breathlessness in people with advanced chronic respiratory disease.

**Methods:** A multicentre, mixed-methods, non-randomised interventional feasibility study with a parallel usual-care control group. Participants were adults with chronic respiratory disease and significant anxiety and breathlessness symptoms (assessed via Integrated Palliative Outcome Scale) receiving care from hospice services. The intervention groups used Alpha-Stim AID for eight weeks either at a fixed or personalised dose, followed by a four-week follow-up period. This feasibility study was not powered to assess clinical efficacy.

**Results:** 12.5% of screened patients at the primary site were eligible, and 29 unique participants were recruited. Three participants withdrew from the study (10.3%), none of which were attributable to CES. Most adverse events were mild, with headache reported frequently across control and intervention groups. Outcome measure completion was high, with data missingness below 6.4%. Numerical rating scales of anxiety and breathlessness fluctuated daily and remained broadly static, whereas GAD-7 scores assessing anxiety improved over time across all groups. This feasibility study was not powered to assess clinical efficacy.

**Conclusion:** CES using Alpha-Stim AID was deliverable within hospice services and was generally acceptable and tolerable among participants who enrolled. Our findings support further evaluation which should involve a fully-powered randomised controlled trial against a sham device to determine whether CES provides clinically meaningful improvements in anxiety for this patient population.

**What is already known on this topic:**

- Anxiety is common amongst patients with chronic respiratory diseases, such as COPD and interstitial lung disease. Management is primarily non-pharmacological.
- Cranial electrotherapy stimulation is used for treatment of anxiety related to primary anxiety disorders, such as generalised anxiety disorder.
- No studies have been carried out exploring cranial electrotherapy stimulation in a palliative care population.

**What this study adds:**

- Cranial electrotherapy stimulation using Alpha-Stim AID was deliverable within hospice services. The intervention was generally accepted and tolerable amongst enrolled participants.
- Headache was common across all three groups. Dizziness was more commonly reported in the personalised dose group.

**How this study might affect research, practice or policy:**

- Our findings support the future development of a definitive randomised controlled trial against a sham device to determine whether CES has a clinically meaningful effect for anxiety in this patient group.

## Introduction

Anxiety is common amongst patients with chronic respiratory diseases, such as chronic obstructive pulmonary disease (COPD) and interstitial lung diseases (ILD).^1, 2^ Prevalence estimates vary according to population and measurement method, but recent systematic reviews report anxiety in approximately 36% of people with COPD and 25% of those with ILD.^3, 4^ Anxiety is tightly interlinked with the experience of breathlessness, and may manifest physiologically as tremor, palpitations, or dyspnoea, making it difficult to distinguish from the symptoms of chronic respiratory disease.^5-7^ Hence, clinically significant anxiety in COPD and ILD often goes under-recognised and unaddressed.^8^ Furthermore, patients with high levels of anxiety experience more frequent exacerbations, which are more likely to result in hospitalisation.^9-11^

Studies evaluating the efficacy of pharmacological interventions (including selective serotonin reuptake inhibitors (SSRIs), tricyclic antidepressants, and azapirones) for anxiety in COPD have shown limited evidence of efficacy in respiratory disease specific presentations.^12^ The mainstay of managing anxiety in chronic respiratory disease is therefore non-pharmacological, including psychological therapies, mind-body interventions, and multidisciplinary care.^13-20^ Pulmonary rehabilitation (a multi-disciplinary programme incorporating education, tailored exercise, and behavioural change interventions) has shown moderate benefit for anxiety symptoms in COPD.^21^ However, evidence is scarce and low-quality for ILD,^22^ and patients with high anxiety are more likely to discontinue pulmonary rehabilitation prematurely due to a lack of perceived benefits.^23, 24^ Patients with advanced disease may find holistic breathlessness services more accessible, which have been shown to reduce distress and may improve psychological outcomes of anxiety and depression.^25^

Cranial electrotherapy stimulation (CES) may offer promise as an additional non-pharmacological management tool as part of holistic breathlessness services offered by specialist palliative care and respiratory services. CES is a non-invasive neuromodulation technique, similar to other electrical non-invasive brain stimulation (NIBS) like transcranial direct current stimulation (tDCS).^26^ Proposed mechanisms include an increase in alpha-oscillatory brain activity (a feature of relaxed wakefulness)^27^ and through stimulation of the vagal nerve.^28^ Alpha-Stim is a CES device which acts to modulate the brain’s electrical activity by delivering a microcurrent of electricity via bilateral electrodes. The Alpha-Stim Anxiety Insomnia and Depression (AID) device is the latest iteration of this device, receiving a CE-mark as a class IIa medical device in 2012. The Alpha-Stim AID device is approximately the size of a mobile phone, battery-powered, and can be used daily in the patient’s home for 20-60 minutes via electrodes attached to the earlobes.

CES treatment can be used alongside pharmacological and psychotherapy treatment, or as a standalone alternative treatment.^29^ A recent general population study found improvements in sleep quality, insomnia symptoms, daytime sleepiness, stress levels, self-efficacy, wellbeing and quality of life.^30^ CES can be delivered through the NHS and reduce anxiety and depression for adults with GAD symptoms.^31^ CES has not been explored for anxiety related to breathlessness, nor in a specialist palliative care setting.

The primary aim of this study was to explore the acceptability and tolerability of CES as a potential intervention for anxiety associated with breathlessness in people with advanced chronic respiratory disease. Secondary objectives were to describe recruitment, retention, adherence, adverse effects, outcome-measure completion and changes in patient-reported outcomes, and to use these findings to inform the design of a future trial.

## Methods

### Study design

This was a multicentre, mixed-methods, non-randomised interventional feasibility study with a parallel usual-care control group. Participants were allocated, as far as possible, according to their preference for CES or usual care. Participants receiving CES continued to receive usual hospice care and used the intervention for eight weeks, followed by a four-week post-intervention follow-up. The study was designed to explore the acceptability and tolerability and to inform the design of a future study of clinical effectiveness.

The study was conducted across three hospices. Ethical approval was granted by the Brighton and Sussex NHS REC (reference 23/LO/0276). This study was registered with clinicaltrials.gov (ID: NCT06066658).

### Participants

Potential participants were identified and initially screened by members of their clinical teams. At the lead site, clinicians maintained a log of screened patients to establish the proportion of outpatients who were eligible for study inclusion.

Participants were eligible if they were 18 years or older, diagnosed with a chronic respiratory disease (including COPD, ILD of any aetiology, or bronchiectasis) with a score ≥2 in the International Palliative Outcome Scale (IPOS)^32^ domains on both ‘shortness of breath’ and ‘anxious or worried’, and a clinician-estimated prognosis of at least 3 months.

Patients were excluded if they had a pacemaker or implantable cardiac defibrillator, or any neurological condition (i.e. epilepsy, brain neoplasm, neurodegenerative disorder, or recent history of brain surgery or a cerebrovascular event in the last 12 months) due to theoretical safety concerns. Patients were also excluded if they had a current or previous episode of psychosis or mania, any current substance abuse or dependence due to theoretical risk of exacerbation, or if pregnant/planning a pregnancy due to unknown potential effects.

Potentially eligible patients completed a consent-to-contact form. A member of the research team then approached the patients to offer a participant information sheet. Where recent IPOS scores were unavailable, these were completed after consent to confirm eligibility.

All patients provided written informed consent.

### Intervention and study procedures

Participants were allocated, as far as possible, according to their preference for either CES plus usual care or usual care alone. Participants allocated to CES received an Alpha-Stim AID device and were trained in its use by a research nurse.

Participants in the initial CES cohort were instructed to use the device at a standard low-dose setting of 100 μA for 60 minutes once daily at home for eight weeks. Tolerability was reviewed on days three and five. Where required because of adverse effects, the current could be reduced to 50 μA and, if necessary, treatment divided into two 30-minute sessions. Participants whose regimen was altered on day five received a further review on day seven. The low-dose starting regimen and predefined adjustments were selected because CES had not previously been studied in this clinically frail population. Participants continued to receive usual hospice care and were followed for a further four weeks after completing the intervention.

The control group received 12 weeks of usual care from their hospice teams and completed the same schedule of assessments.

Following review of early tolerability data and approval of a protocol amendment, a subsequent personalised-dose CES cohort was introduced. Participants increased the current until they experienced mild vertigo and then reduced the setting by 50 μA to establish an individually tolerated dose. All other intervention, follow-up and data-collection procedures were unchanged. Transient vertigo occurring during dose titration was regarded as an expected part of the titration procedure and was not recorded as an adverse event; any recurrent vertigo after the individualised dose had been established was recorded as an adverse event.

### Outcome measures

Feasibility outcomes were measured by recruitment and retention rates, adherence to the device (intervention group only), frequency and nature of adverse events (AEs), and completeness of outcome data. Regarding AEs, these were monitored throughout the 12-week study period. At each study contact participants were asked about symptoms previously associated with Alpha-Stim use: headache, dizziness, vertigo, nausea, tinnitus and skin irritation. Each event was assessed for causality (table 2) and severity by a senior palliative care clinician. Causality assessment was intentionally conservative: events for which causality could not definitively be excluded were classified as ‘possible’ or ‘probably related.

Patient-reported outcome measures included several questionnaires, including the Generalised Anxiety Disorder-7 (GAD-7), Patient Health Questionnaire-9 (PHQ-9), Sleep Condition Indicator (SCI), EuroQol’s health-related quality of life measure: EQ-5D-5L, Dyspnoea-12 (D-12), Chronic Respiratory Questionnaire (CRQ-SR), and Integrated Palliative Outcome Scale (IPOS). These were completed at baseline and weeks 2 (except IPOS), 4, 8 and 12 by all participants.

Participants in all groups also completed a daily diary, which included a numerical rating scale (NRS) from 0-10 for anxiety (at worst, at best, and on average) and breathlessness (at worst, at best, and on average).

### Qualitative methods

All participants were also invited to take part in a single semi-structured interview with a qualitative researcher at the end of week 12. The study patient and public involvement group (PPI) supported the development of a topic guide which explored participants’ experiences of their anxiety and breathlessness, using the Alpha-Stim AID (intervention groups only), and being involved in the study.

### Sample size

As this was a feasibility study, no formal sample size calculation was carried out. The original target was for 10 participants to complete each of the fixed-dose CES and usual-care control groups. Following the protocol amendment introducing the personalised-dose CES cohort, the overall target was increased to 30 completed study entries, with 10 in each group. Recruitment ended once this had been met.

### Statistical methods

Analyses were primarily descriptive and carried out to assess recruitment, retention, adherence, adverse events, and completeness of outcome data rather than to evaluate clinical efficacy. Baseline characteristics were summarised using means and standard deviations, or frequencies and percentages as appropriate. Outcome measures were summarised descriptively at each time point using available case analysis to handle missing data. Associations between daily NRS-Breathlessness and NRS-Anxiety were assessed using Pearson’s correlation coefficient.^33^

## Results

### Recruitment

A total of 29 unique patients were recruited. Four participants who completed the fixed-dose cohort subsequently re-entered the personalised dose arm, for a total of 33 study entries. At the main site, 12.5% of screened patients met eligibility criteria. Figure 1 illustrates patient flow.

**Figure 1:**
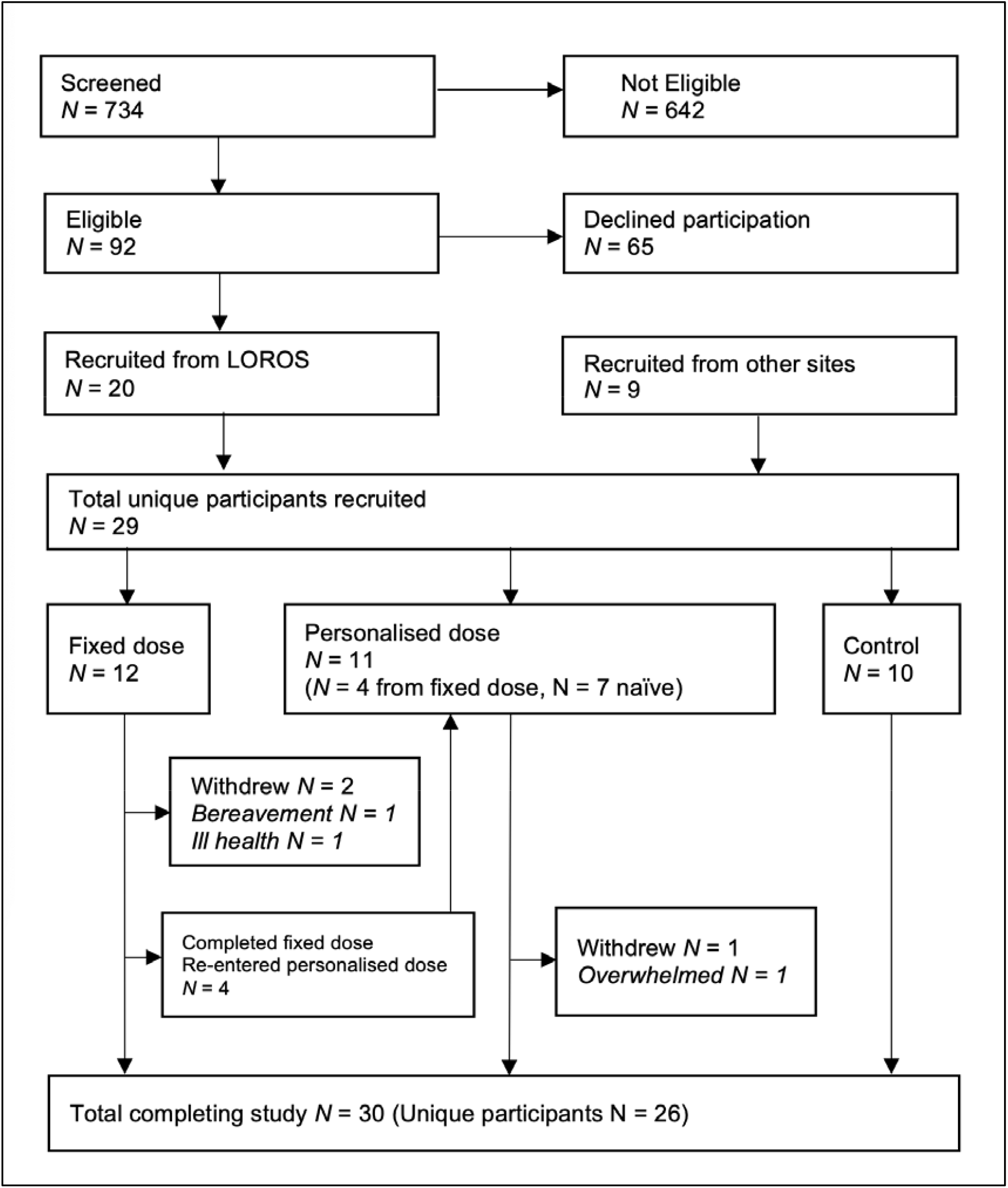
Participant flow diagram.

Three participants withdrew: two from the fixed dose arm, and one from the personalised dose arm. One withdrew due to bereavement (week 2), one withdrew due to ill health (week 8), and one felt the study was too overwhelming to complete (week 3).

### Study cohort

Baseline characteristics are shown in Table 1. COPD was the most common primary diagnosis, followed by ILD. Most enrolled patients had severe symptoms at point of enrolment (IPOS of 3 across both domains).

**Table 1:** Baseline Characteristics.

| Characteristic* | Total Unique Participants | Control | Alpha Stim (Fixed Dose) | Alpha Stim (Personalised Dose) |
| --- | --- | --- | --- | --- |
|  | N = 29 | N = 10 | N = 12 | N = 11 |
| Sex |  |  |  |  |
| **Female** | 15 (51.7%) | 6 (60%) | 7 (58.3%) | 3 (27.3%) |
| **Male** | 14 (48.3%) | 4 (40%) | 5 (41.7%) | 8 (72.7%) |
| **Primary diagnosis** |  |  |  |  |
| **COPD** | 14 (48.3%) | 5 (50%) | 5 (41.7%) | 7 (63.6%) |
| **ILD** | 10 (34.5%) | 3 (30%) | 4 (33.3%) | 3 (27.3%) |
| **Other** | 5 (17.2%) | 2 (20%) | 3 (25%) | 1 (9.1%) |
| **Main ethnic group** |  |  |  |  |
| **Asian/British Asian** | 4 (13.8%) | 1 (10%) | 3 (25%) | 1 (9.1%) |
| **White** | 25 (86.2%) | 9 (90%) | 9 (75%) | 10 (90.9%) |
| **Age, years mean (std dev)** | 65.6 (8.0) | 67 (7.7) | 64 (8.7) | 67 (7.9) |
| **Baseline IPOS score median (IQR)** |  |  |  |  |
| **Shortness of breath** | 3 (3, 3) | 3 (2, 3) | 3 (3, 4) | 3 (3, 3) |
| **Anxious or worried** | 3 (2, 4) | 2.5 (2, 3) | 3.5 (2, 4) | 3 (2, 4) |
| **\*Values expressed as N(%) unless otherwise indicated** |  |  |  |  |

### Adherence

Participants who completed the full study period were should have used CES for 60 minutes per day, for a total of 56 hours. Due to device limitations, usage time was only recorded from day 3 onwards; therefore, the maximum recorded usage was 53 hours, which was treated as 100% of measurable prescribed use.

In the fixed regime group participants self-reported 54 hours and 3 minutes of use on average, whereas device interrogation showed 39 hours and 48 minutes. Participants reported missed sessions descriptively in the daily log as feeling unwell, being interrupted, or device issues i.e. batteries running out.

In the personalised dose group participants self-reported 46 hours and 50 minutes of use on average, whereas device interrogation showed 39 hours and 13 minutes. Participants reported non-adherence due to feeling unwell, forgot, the ear clip broke, they were too busy, or they had sore ears.

### Adverse events

Most events across all groups were mild severity (78.1% of control AEs, 79.6% of fixed dose AEs, and 90.0% of personalised dose AEs). The causality of AE to the study intervention is shown in Table 2, and the nature of AEs is shown in Table 3.

**Table 1:** Causality of adverse events across both Alpha-Stim groups.

|  | **Fixed Dose** | **Personalised Dose** |
| --- | --- | --- |
| **Causality** | **Frequency (%)** | **Frequency (%)** |
| **Definitely Related** | 13 (9.5%) | 13 (6.7%) |
| **Probably Related** | 40 (29.2%) | 100 (51.3%) |
| **Possibly Related** | 31 (22.6%) | 51 (26.1%) |
| **Not Related** | 53 (38.7%) | 31 (15.9%) |

**Table 2:** Outcome measures across 12-week study period.

|  | Timepoint | Number of responses | Control group<br>N=10 | Fixed dose group<br>N=12<br>Mean (95% CI) | Personalised dose<br>group N=11 |
| --- | --- | --- | --- | --- | --- |
| **GAD-7** | Baseline | 33 | 12.1 (7.7, 16.5) | 13.4 (9.5, 17.4) | 12 (8.4, 15.6) |
|  | Week 2 | 30 | 10.9 (6.7, 15.1) | 9.6 (6.9, 12.4) | 9.1 (4.2, 14) |
|  | Week 4 | 29 | 10.2 (6, 14.4) | 9.5 (6.4, 12.7) | 10.1 (6.2, 14) |
|  | Week 8 | 28 | 9 (5, 13) | 8.8 (5.1, 12.5) | 7.9 (3.7, 12.1) |
|  | Week 12 | 29 | 8.6 (4.6, 12.6) | 8.5 (5.3, 11.7) | 8.1 (3.7, 12.5) |
| **CRQ-SR Emotional** | Baseline | 33 | 23.6 (19.3, 27.9) | 20.1 (15.4, 24.7) | 20.8 (17, 24.7) |
|  | Week 2 | 30 | 23.6 (17.7, 29.5) | 23.5 (20.9, 26.2) | 22 (15.8, 28.2) |
|  | Week 4 | 31 | 23.6 (17.4, 29.8) | 24.5 (20.3, 28.6) | 24.6 (20.1, 29.1) |
|  | Week 8 | 28 | 24.9 (19.3, 30.5) | 26.1 (19.8, 32.4) | 24.6 (19.5, 29.6) |
|  | Week 12 | 30 | 25.9 (21.5, 30.3) | 26 (20.6, 31.4) | 21.5 (15.7, 27.3) |
| **CRQ-SR Mastery** | Baseline | 33 | 12.9 (9.6, 16.2) | 10.1 (7.3, 12.8) | 11.5 (9.9, 15.9) |
|  | Week 2 | 30 | 12.1 (8.1, 16.1) | 12.5 (9.9, 15.1) | 12.9 (9.9, 15.9) |
|  | Week 4 | 31 | 12.7 (8.6, 16.8) | 13.4 (11.2, 15.6) | 12.9 (10.7, 15.1) |
|  | Week 8 | 28 | 12.9 (9.3, 16.5) | 12.7 (9.4, 16) | 12.3 (9.8, 14.8) |
|  | Week 12 | 30 | 12.9 (10, 15.8) | 13.4 (10.2, 16.6) | 12.7 (9.5, 15.9) |
| **CRQ-SR Dyspnoea** | Baseline | 33 | 9.7 (7.5, 11.9) | 8.8 (6.1, 11.6) | 9.6 (8.2, 11.1) |
|  | Week 2 | 30 | 9.7 (6.3, 13.1) | 9.2 (7, 11.4) | 9.6 (7.2, 12) |
|  | Week 4 | 31 | 10.7 (7.3, 14.1) | 10.2 (7.6, 12.8) | 8.7 (6.5, 10.9) |
|  | Week 8 | 28 | 11 (8, 14) | 10 (6.7, 13.3) | 11.1 (6.8, 15.5) |
|  | Week 12 | 30 | 9.7 (6.8, 12.6) | 9.6 (6.8, 12.4) | 9.8 (6.8, 12.8) |
| **CRQ-SR Fatigue** | Baseline | 33 | 11 (8.3, 13.7) | 8.5 (6.9, 10.1) | 9.5 (7.6, 11.5) |
|  | Week 2 | 30 | 11.1 (8.1, 14.1) | 10.3 (7.8, 12.8) | 10.9 (8.4, 13.4) |
|  | Week 4 | 31 | 11.6 (9.7, 13.5) | 11.5 (9.3, 13.6) | 11.2 (9, 13.4) |
|  | Week 8 | 28 | 11.3 (8.8, 13.9) | 12.1 (8.6, 15.6) | 10.7 (8.6, 12.7) |
|  | Week 12 | 30 | 10.9 (8, 13.8) | 11.3 (8.9, 13.7) | 10.6 (7.8, 13.4) |
| **D-12** | Baseline | 33 | 22.2 (18.1, 26.3) | 24.8 (19.4, 30.1) | 25.3 (20, 30.6) |
|  | Week 2 | 30 | 23.5 (16.4, 30.6) | 24.1 (19.9, 28.3) | 25 (19.6, 30.4) |
|  | Week 4 | 31 | 23.3 (18.3, 28.3) | 20.9 (17, 24.9) | 25.6 (21.8, 29.4) |
|  | Week 8 | 28 | 21.8 (18.7, 24.9) | 19.7 (15, 24.4) | 24.9 (19.8, 30) |
|  | Week 12 | 30 | 20.9 (17.6, 24.2) | 17.3 (13.5, 21.1) | 24.1 (18.9, 29.3) |
| **PHQ-9** | Baseline | 33 | 11.1 (6.9, 15.3) | 16.9 (13.7, 20.1) | 13.9 (10.4, 17.4) |
|  | Week 2 | 30 | 12.7 (8.1, 17.3) | 14.4 (11, 17.7) | 14 (8.7, 19.3) |
|  | Week 4 | 31 | 11 (6.2, 15.8) | 10.4 (7.5, 13.2) | 10 (5.2, 14.8) |
|  | Week 8 | 28 | 9.7 (6.2, 13.1) | 9.9 (5.9, 13.9) | 10 (5.4, 14.6) |
|  | Week 12 | 30 | 10.8 (6.7, 14.9) | 10.3 (6.6, 14) | 8.3 (4.9, 11.7) |
| **SCI** | Baseline | 30 | 12.4 (8, 16.8) | 13.6 (6.6, 20.6) | 14.8 (7.8, 21.8) |
|  | Week 2 | 30 | 13.8 (8.9, 18.7) | 13.5 (7, 20) | 14 (7.9, 20.1) |
|  | Week 4 | 30 | 14.8 (9.7, 19.9) | 14.8 (8.6, 21) | 18.8 (11.7, 25.8) |
|  | Week 8 | 27 | 12.9 (8, 17.8) | 17.2 (9.5, 24.9) | 20.4 (14.6, 26.2) |
|  | Week 12 | 28 | 12.8 (8.8, 16.8) | 16.9 (9.2, 24.6) | 18.2 (11.9, 24.5) |
| **IPOS: How has shortness of breath affected you?** | Baseline | 33 | 2.7 (2.4, 3) | 3.3 (3, 3.6) | 3 (2.7, 3.3) |
|  | Week 4 | 31 | 2.7 (2.2, 3.2) | 2.6 (2.3, 3) | 2.7 (2, 3.4) |
|  | Week 8 | 28 | 2.8 (2.1, 3.4) | 2.2 (1.4, 3) | 2.8 (2.1, 3.4) |
|  | Week 12 | 30 | 2.7 (2, 3.4) | 2.7 (2.1, 3.3) | 2.6 (2.2, 3) |
| **IPOS: Have you been feeling anxious or worried about your illness or treatment?** | Baseline | 33 | 2.6 (2.1, 3.1) | 3.2 (2.6, 3.8) | 3 (2.5, 3.5) |
|  | Week 4 | 31 | 2.7 (2.1, 3.3) | 2.5 (1.8, 3.1) | 2.7 (1.7, 3.7) |
|  | Week 8 | 28 | 2.2 (1.4, 3.1) | 2.3 (1.3, 3.3) | 2.9 (1.9, 3.9) |
|  | Week 12 | 30 | 2.3 (1.8, 2.8) | 2.2 (1.2, 3.2) | 2.7 (2.2, 3.2) |
| **EQ-5D-5L** | Baseline | 33 | 48.5 (34.2, 62.8) | 47.1 (36, 58.1) | 53.6 (40.4, 66.8) |
|  | Week 2 | 30 | 47 (33, 61) | 49.6 (39.2, 60.1) | 43.1 (25.2, 61) |
|  | Week 4 | 31 | 39.8 (25.2, 54.4) | 53.8 (38.9, 68.7) | 61.8 (47, 76.6) |
|  | Week 8 | 28 | 45.6 (28.3, 62.9) | 56.1 (45.5, 66.7) | 57.6 (48.4, 66.7) |
|  | Week 12 | 29 | 41.3 (22.2, 60.5) | 56.5 (43.7, 69.3) | 54.9 (42.1, 67.7) |

**Table 3:** Nature of adverse events.

| Adverse event | Number of adverse events (percentage) |  |  |
| --- | --- | --- | --- |
|  | Control group<br>N = 10 | Fixed dose<br>N = 12 | Personalised dose<br>N = 11 |
| Headache | 24 (32.9%) | 51 (37.2%) | 69 (35.4%) |
| Dizziness or light-headedness | 9 (12.3%) | 15 (10.9%) | 42 (21.5%) |
| Tinnitus | 6 (8.2%) | 8 (5.8%) | 16 (8.2%) |
| Nausea | 5 (6.8%) | 7 (5.1%) | 2 (1.0%) |
| Tingling sensation | 0 | 7 (5.1%) | 4 (2.1%) |
| Skin irritation | 0 | 8 (5.8%) | 1 (0.05%) |
| Other | 29 (34.3%) | 41 (26.2%) | 61 (31.3%) |
| **TOTAL** | **73** | **137** | **195** |

‘Other’ AEs were primarily due to intercurrent illness or exacerbation of their respiratory condition (e.g. respiratory tract infections, pyrexia, palpitations, diarrhoea and vomiting, or abdominal pain). There were no serious adverse events.

### Completion of study documents

Across the 12-week study period, missing data were generally low in the daily logs. In the control group, missing values were 0.97% (n=7, where n=missing data points), 2.35% (n=17) and 0.86% (n=6) respectively for best, average and worst breathlessness, and 3.18% (n=23), 4.15% (n=30) and 3.04% (n=22) respectively for best, average and worst anxiety.

In the fixed dose group, missing values were higher at 3.81% (n=28), 6.4% (n=47) and 3.81% (n=28) respectively for best, average and worst breathlessness, and 4.09% (n=30), 6.13% (n=45) and 4.09% (n=30) respectively for best, average and worst anxiety, with similar figures in the personalised dose group.

Missing data for the questionnaires were also generally low at 2.35% (n=82) for the fixed dose, 4.46% (n=145) for the personalised dose, and 2.46% (n=70) for the control group. Across all groups, reasons for missing data were recorded as too unwell to complete questionnaires (64.0%), questionnaires not returned to research nurse (20.9%), question unintentionally missed by participant (8.6%), participant felt unable to answer the question (5.7%), or question too upsetting (0.7%). This related to an element of the CRQ-SR which asks participants to identify activities that cause them to feel breathless.

### NRS-Breathlessness and NRS-Anxiety

NRS-Breathlessness and NRS-Anxiety scores fluctuated day-to-day in all three groups. Minimal change was observed from baseline to week 12, although this study is not powered to investigate clinical effect. Across all groups, participants varied in how far apart they ranked best, average, and worst symptoms. Best, average, and worst NRS scores are shown for breathlessness in Figure 2 and anxiety in Figure 3.

**Figure 2:**
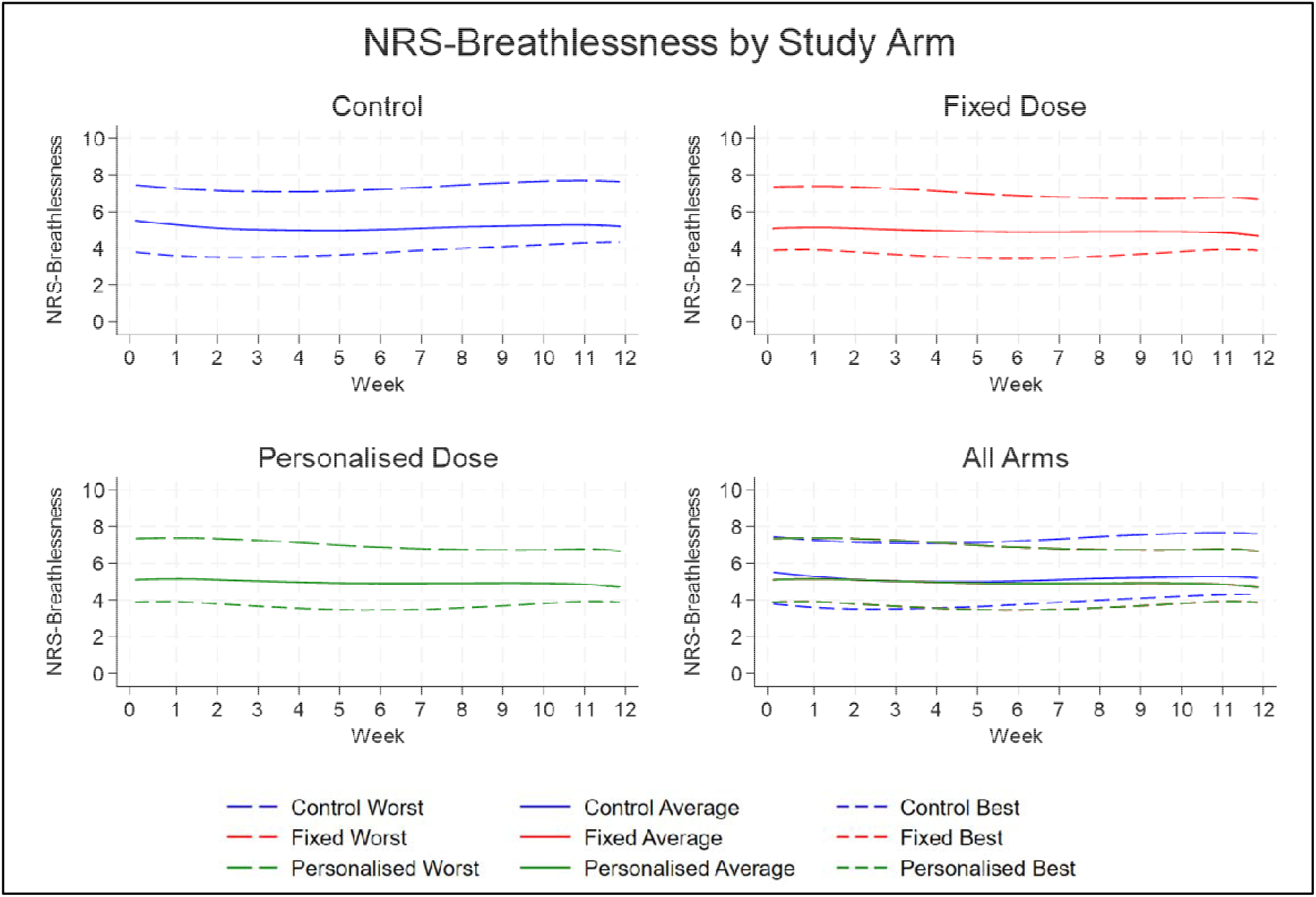
NRS-Breathlessness by Study Arm.

**Figure 3:**
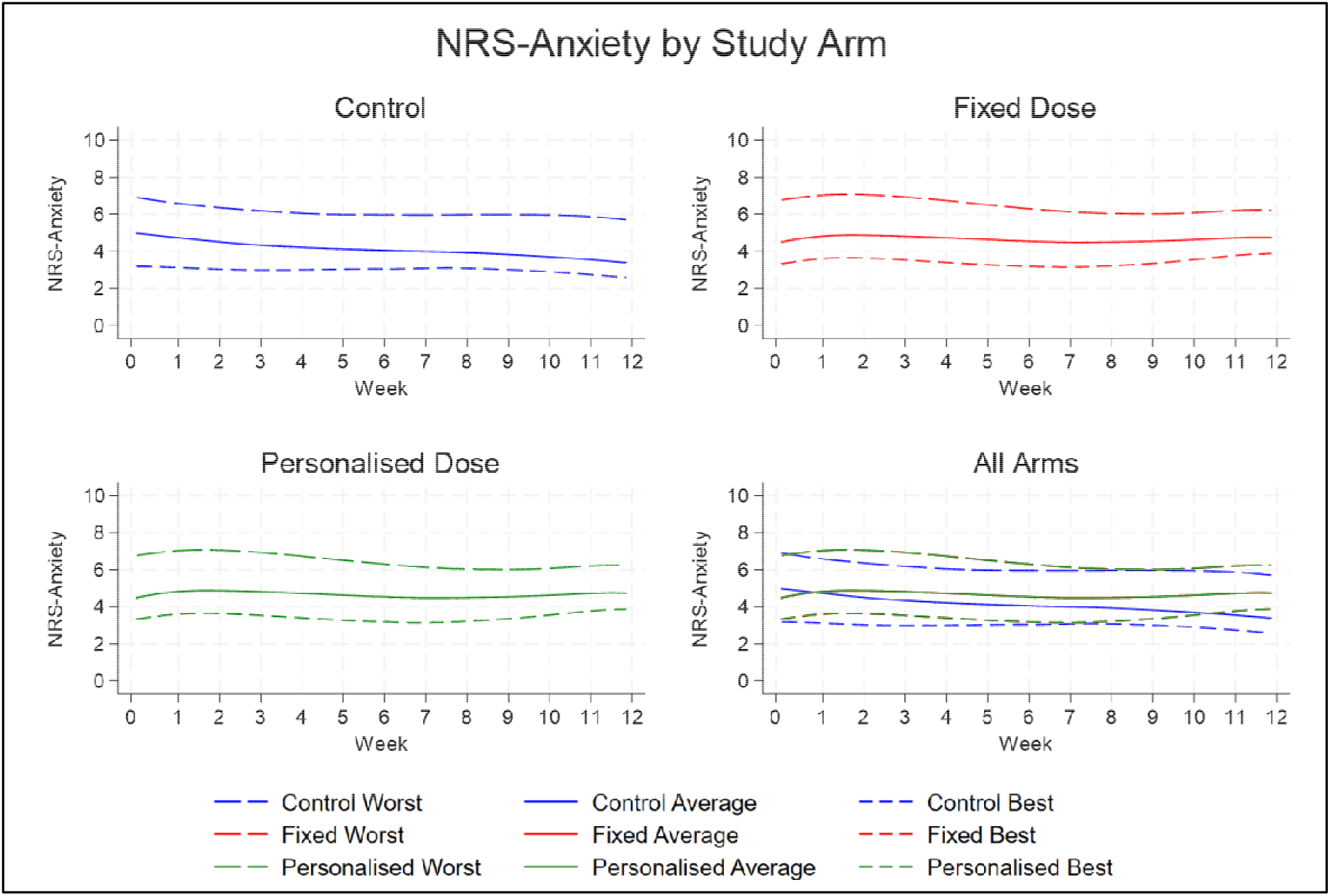
NRS-Anxiety by Study Arm.

The linear association between anxiety and breathlessness varied across groups. In the control group, correlations were of fair strength (r = 0.35-0.50), whereas the fixed dose group showed moderately strong correlation (r = 0.70-0.78). The personalised dose group showed poor to fair correlations (r = 0.28-0.38), indicating a weaker association between anxiety and breathlessness in this cohort.

### Patient-reported outcome measures

Baseline GAD-7 scores indicated moderate anxiety in all groups, with mean values of 12.1 (7.7, 16.5) in the control group, 13.4 (9.5, 17.4) in the fixed-dose group, and 12.0 (8.4, 15.6) in the personalised dose group. Scores decreased across the 12-week study period in all groups, with all groups in the mild anxiety range at week 12, as shown in Table 4.

Across the remaining outcome measures, baseline scores were broadly similar between groups. Modest improvements over time were observed for CRQ-SR emotional and mastery domains, depressive symptoms (PHQ-9), sleep (SCI), and IPOS anxiety scores. Dyspnea-12 and IPOS breathlessness scores showed some reduction in breathlessness, particularly in the fixed-dose group, while EQ-5D-5L scores indicated stable or improved quality of life in the intervention groups. However, 95% confidence intervals were wide and overlapped substantially between treatment groups and across timepoints, including between baseline and week 12, with no clear separation between treatment arms. These findings are descriptive only and should not be interpreted as evidence of treatment effect in this small sample.

## Discussion

### Principal findings and interpretation

The aim of this feasibility study was to assess the acceptability and tolerability of CES, and to inform whether progression to a subsequent definitive trial would be justified. The findings should therefore be interpreted primarily in relation to feasibility rather than efficacy. Although outcome data were collected across various symptom domains, this study was not powered to detect differences between groups or estimate clinical effect. As no formal progression thresholds were prespecified, feasibility was interpreted collectively across recruitment, retention, adherence, tolerability and the completeness of the outcome data.

### Recruitment and retention

Our eligibility-to-enrolment pathway at the primary hospice site demonstrated 12.5% of screened patients were eligible. Of those eligible 20/92 (21.8%) consented to enrol in the study. This eligibility to enrolment conversion rate supports the acceptability of the study concept, including a willingness to consider a device-based intervention for anxiety related to breathlessness. However, our recruitment period required extension to achieve recruitment targets, as enrolment to the study was slower than anticipated.

Recruitment challenges are well recognised in clinical trials across palliative care populations for several reasons, including fluctuating health, high symptom burden, and competing clinical priorities.^34^ It is likely that all these factors affected our patient population to varying degrees. Whilst observed enrolment rate provides support for acceptability, the overall screening-to-enrolment pathway highlights the need for careful consideration of recruitment pathways for any future definitive trial.

Three of the 29 unique participants withdrew, representing a withdrawal rate of 10.3%. Although our withdrawal rate is higher than studies using CES in psychiatric populations (2.1%),^35^ it is substantially lower than other studies of participants with significant physical illness. For example, Ree et al (2024) demonstrated an attrition rate of 33% in their study of adults with fibromyalgia using CES.^36^ The relatively low withdrawal rate in this medically frail population provides encouraging evidence that the intervention and study procedures were generally tolerable to those who enrolled.

### Outcome measure acceptability

Rates of missing data were generally low despite the substantial volume of outcome measures participants were asked to complete, and feedback indicated that the questionnaires were generally well tolerated. One questionnaire item was reported as being too upsetting to complete by one participant. Whilst this was a single occurrence, it highlights the potential for emotional impact with some patient-reported outcome measures.

### Adverse events and tolerability

Although the frequency of reported AEs, particularly headache, was higher than has been described in other CES research, a similarly high incidence of headache was also observed in the control group.^35^ Patients with chronic respiratory disease are likely to experience troublesome headaches as a manifestation of chronic hypoxaemia; research within COPD populations demonstrate a prevalence of 31.9%.^37^ This is remarkably similar to the incidence rate of headaches in our study, suggesting this is likely reflective the symptom burden in this patient population than attributable to the intervention. Most prior CES work has been conducted in younger populations with primary psychiatric disorders rather than in a specialist palliative care population with significant physical illness, therefore direct comparisons of adverse event frequencies between studies should be interpreted with caution.

AEs were more frequently reported in the personalised dose group, particularly dizziness. Although these AEs were more commonly reported, the overall tolerance is supported by the absence of withdrawals from the personalised dose group. For future studies, reporting AEs in relation to phase of treatment (i.e. during the initial phases of dose titration versus maintenance later in the study) would clarify if these symptoms are an expected consequence of establishing a dose or persistent effects.

The high level of adherence amongst both the fixed dose and personalised dose groups is encouraging, with participants completing most daily treatment sessions. This demonstrates that CES is an accessible treatment option even in the context of high symptom burden and chronic disease requiring specialist palliative care input. High adherence is consistent with other CES research.^35^

### Integrated Findings

Table 5 integrates both the quantitative findings from this report and the qualitative findings outlined in a separate publication.^38^ The integrated analysis demonstrates convergence across most of the feasibility outcomes with suggestions for improvement in a definitive trial. Discordance was demonstrated regarding preliminary clinical outcomes (GAD-7 and NRS). Qualitative data may demonstrate a social desirability bias, whereby the participants wanted to convey a feeling of effect to the researcher. Our selected outcome measures may not be sensitive enough to detect the benefits our participants describe in the qualitative data, as these primarily focused on symptom severity whilst participants described more nuanced benefits around coping and emotional wellbeing. Ultimately, this study was not designed or powered to detect efficacy, therefore conclusions around clinical effect should not be drawn from this data.

**Table 5:** Integration of quantitative and qualitative findings.

| **Feasibility domain** | **Quantitative finding** | **Qualitative finding** | **Integrated interpretation** |
| --- | --- | --- | --- |
| **Recruitment** | 745 screened; 95 eligible; 29 unique participants enrolled; 6 post-consent exclusions due to IPOS <2; recruitment period extended to reach target. | Participants described anxiety related to breathlessness as an important unmet need; PPI work indicated a 'just try anything' mindset. | Recruitment was feasible but slower than anticipated. Eligibility assessment using IPOS after consent created avoidable inefficiency and should be revised. |
| **Retention** | 3/29 enrolled participants withdrew after enrolment. | Reasons included advancing illness, bereavement, and feeling overwhelmed. | Attrition was low overall, but study burden may need reducing for some participants. |
| **Intervention acceptability** | 10 participants in each group completed the intervention; no withdrawals due to device AEs. | Participants described the device as simple to understand and easy to use. | Concordance between quantitative and qualitative data suggesting acceptability of intervention. |
| **Adherence/ device use** | Self-reported adherence higher than via device interrogation. | Participants incorporated device into daily routine, primarily evenings; challenges included illness and occasional technical issues. | Reasonable adherence suggested. Definitive trial should consider recording adherence via device interrogation rather than self-reported measures for accuracy. |
| **Outcome completion** | 26/29 completed baseline and all follow-up visits; data missingness generally low; | Participants described questionnaires as manageable with no preference for which questionnaire | Outcome collection was feasible, although questionnaire burden could be streamlined. |
|  | missing data primarily due to ill health. | represented their experience best. |  |
| **Safety** | 405 adverse events; no serious adverse events; no device-related SAEs. | Participants reported occasional tolerable and transient side effects, usually eliminated by adjusting dose. | Both datasets support tolerability. High incidence of headache likely to reflect underlying physical illness. |
| **Preliminary clinical outcomes** | Mean change of 4.1 points in the GAD-7 across all groups; static NRS scores for both anxiety and breathlessness. | Participants reported subjective experience of positive change, particularly regarding positive mindset, ability to cope, and enhanced emotional wellbeing. | Discordance between quantitative and qualitative findings, and across outcome measures. |

### Limitations

This study has several limitations. The non-randomised design may have introduced baseline differences between groups and limits interpretation of between group comparisons. This design was informed by PPI work, which indicated a small minority of patients held strong preferences around not using a device that incorporated an electrical current. The study was conducted within specialist hospice and breathlessness services, which may limit generalisability to people with less advanced disease or those outside palliative-care settings. A further limitation is the repeated participation by a small number of participants in both the fixed dose and the personalised dose arms which would not be possible in a definitive trial. Though a four-week washout period was considered sufficient, this may have influenced participants familiarity and expectations of the intervention. Device-recorded adherence was limited to cumulative usage time and could not confirm the timing, duration or number of treatment sessions.

### Implications

This is the first study to evaluate CES specifically for anxiety associated with breathlessness, whereas previous research investigating CES has largely focused on primary anxiety disorder or depression. The qualitative data from this study suggests that participants frequently described how using the Alpha-Stim AID provided a structured opportunity to relax and regain a sense of control.^38^ The perceived benefits of CES may therefore originate from not only neuromodulatory effects, but an established daily routine encouraging relaxation and symptom self-management. Future research therefore should include a randomised controlled trial using a sham device to establish clinical efficacy and understand the true mechanism of action.

## Conclusion

CES using Alpha-Stim AID was deliverable within hospice services, was generally acceptable and tolerable among participants who enrolled. Recruitment, adherence measurement and outcome selection would require refinement before a definitive trial. These findings support further evaluation that should include a fully-powered randomised controlled trial against a sham device to determine whether CES provides clinically meaningful improvements in anxiety for this patient population.

## Contributors

This study was conceptualised by CF, CG and LB. Statistical analysis was carried out by SC and SB, supervised by LG. Qualitative data analysis was carried out by KW, LB and CG. LB drafted the final manuscript with SC, LG and DW. CF was the initial Chief Investigator of the study prior to retirement, followed by DW. All authors have approved the final version to be published.

## Ethical approval

Ethical approval was granted by the Brighton and Sussex NHS REC (reference 23/LO/0276) on 3^rd^ May 2023.

## Data availability statement

Data are available from the corresponding author upon reasonable request. The data generated in this study are subject to patient confidentiality, and the transfer of data or materials will require appropriate institutional data protection approval.

## Acknowledgements

The authors wish to sincerely thank all participants for so generously giving their time to this study. Special thanks to Sue Ashton, Janis Hayward, David Riley, and David Miodrag for their support of this study.

